# Gender differences in the determinants of willingness to get the COVID-19 vaccine among the working-age population in Japan

**DOI:** 10.1101/2021.04.13.21255442

**Authors:** Tomohiro Ishimaru, Makoto Okawara, Hajime Ando, Ayako Hino, Tomohisa Nagata, Seiichiro Tateishi, Mayumi Tsuji, Shinya Matsuda, Yoshihisa Fujino, for the CORoNaWork Project

**Author notes:** Corresponding author: Tomohiro Ishimaru, MD, MPH, PhD, Department of Environmental Epidemiology, Institute of Industrial Ecological Sciences, University of Occupational and Environmental Health, 1-1 Iseigaoka, Yahata-nishi-ku, Kitakyushu, Fukuoka 807-8555, Japan.

## Abstract

Many factors are related to vaccination intentions. However, gender differences in the determinants of intention to get the coronavirus disease 2019 (COVID-19) vaccine have not been fully investigated. This study examined gender differences in the determinants of willingness to get the COVID-19 vaccine among the working-age population in Japan. We conducted a cross-sectional study of Japanese citizens aged 20–65 years using an online self-administered questionnaire in December 2020. Logistic regression analysis was performed. Among 27,036 participants (13,814 men and 13,222 women), the percentage who were willing to get the COVID-19 vaccine was lower among women than among men (33.0% vs. 41.8%). Age and education level showed a gender gap regarding the association with willingness to get the COVID-19 vaccine: men who were older or had a higher level of education were more willing to get the vaccine, whereas women aged 30–49 years and those with a higher level of education showed a relatively low willingness to get the vaccine. For both men and women, marriage, higher annual household income, underlying disease, current smoking, vaccination for influenza during the current season, and fear of COVID-19 transmission were linked to a higher likelihood of being willing to get the COVID-19 vaccine. These findings give important insight into identifying target groups in need of intervention regarding COVID-19 vaccination, especially among women. Providing education about COVID-19 and influenza vaccination in the workplace may be an effective strategy to increase COVID-19 vaccine uptake.

## Introduction

Vaccines are expected to be a key measure against the coronavirus disease 2019 (COVID-19) pandemic.^1^ Vaccine development usually takes decades, and, although a variety of COVID-19 vaccines are currently being developed at an unprecedented rate, evidence has not yet established their long-term safety.^2^ In Japan, the COVID-19 vaccination program has been administered since February 2021, with eligibility proceeding in the following order: healthcare workers, older adults (aged 65 years or older), and people with underlying diseases and caregivers working at care facilities for older adults.^3^ Other people will become eligible for vaccination at a later date, following the populations listed above, which are the main sources of infection.^4^ To reach herd immunity, most citizens need to be vaccinated.^5^

Vaccination intentions are recognized as the most important issue in the rollout of vaccination programs.^6^ However, Japan is known for its lack of public trust in vaccines^7^ because negative campaigns against the vaccination—highlighting scandals and severe adverse effects, for instance—have increased hesitation concerning some vaccinations over the past several decades.^8–11^ For example, a national human papillomavirus vaccination program for girls was launched in 2013 but was discontinued after 3 months because of sensational case reports of adverse effects.^12^ This event had a negative impact on vaccination intentions, especially among women.^13^

Various factors are related to COVID-19 vaccination intentions. Previous studies have reported that age, gender, race, education level, influenza vaccination status, and fear of COVID-19 transmission are associated with the willingness to get the COVID-19 vaccine.^14–18^ In Japan, willingness to get the COVID-19 vaccine was previously examined in a single study of 1,100 adults, with results showing that vaccine hesitancy was higher in Japan than in other countries, particularly among women.^19^ However, gender differences in the determinants of willingness to get the COVID-19 vaccine, such as demographic and health characteristics, influenza vaccination status, and fear of COVID-19 transmission, have not been fully investigated. Furthermore, more studies involving larger samples of the working-age population are needed. Therefore, the current study examined gender differences in the determinants of willingness to get the COVID-19 vaccine among the working-age population in Japan. This study will give important insight into identifying target groups in need of intervention regarding COVID-19 vaccination.

## Materials and methods

### Study design

We conducted a cross-sectional study about COVID-19 among the working-age population in Japan on December 22–26, 2020. The present article is part of a series of studies conducted under the CORoNaWork (Collaborative Online Research on the Novel-coronavirus and Work) Project.^20^ This study was conducted during the third wave of the COVID-19 pandemic in Japan, when the number of infected people and deaths peaked.^21^ Although COVID-19 vaccination began in December 2020 in the United Kingdom (UK) and the United States (US), Japan had not yet begun to administer COVID-19 vaccines during the survey period.^22^ Pfizer requested a fast-track approval of its COVID-19 vaccine from the government of Japan on December 18, 2020 and became the first company to receive this approval on February 14, 2021.^3^

Criteria for eligibility for the present study were being aged 20–65 years and currently working. Healthcare workers and caregivers were not invited to participate. We recruited study participants from the panelists of Cross Marketing Inc. (Tokyo, Japan), an Internet research company. These panelists regularly respond to self-administered Internet surveys distributed by the company, receiving tokens that can be redeemed for products and services as compensation.

For this survey, we used quota sampling by gender, residence (five districts), and occupation (office worker or other). Information on these characteristics was retrieved from the panelists’ Cross Marketing Inc. registration information. Cross Marketing Inc. sent an invitation email to panelists who met the eligibility criteria, and those who were interested in the study proceeded to the survey via a hyperlink. The participants provided informed consent prior to beginning the online questionnaire. We set a quota of 1,650 participants for each of the 20 strata and ceased recruitment when the target number was reached. In total, 33,087 participants from all over Japan completed the questionnaire. After excluding invalid responses, a total of 27,036 participants remained. All participants received standard incentives through Cross Marketing Inc. (i.e., a few dollars’ worth of tokens). The study was conducted according to the guidelines of the Declaration of Helsinki, and it was approved by the Ethics Committee of the University of Occupational and Environmental Health, Japan (Approval number: R2-079).

### Measures

The survey questions included items on demographic and health characteristics, influenza vaccination status during the current season, fear of COVID-19 transmission, and willingness to get the COVID-19 vaccine. The demographic and health characteristics were gender, age, education, marital status, annual household income (1 USD was equal to 106.78 JPY, using 2020 conversion rates),^23^ underlying disease, and smoking status. In line with a previous study,^18^ we developed the following item to assess willingness to get the COVID-19 vaccine: “If a COVID-19 vaccine becomes available, I will get it.” The possible answers to this question were *yes* and *no*. Fear of COVID-19 transmission was assessed by asking “Do you fear COVID-19 transmission?”, with *yes* and *no* as the possible responses.

### Data analysis

We calculated frequencies and proportions for all variables. Descriptive statistics were also calculated by gender to reveal more detailed background factors. Logistic regression analysis was performed to calculate adjusted odds ratios (aORs) and 95% confidence intervals (CIs) for all variables to evaluate associations with willingness to get the COVID-19 vaccine. Because we hypothesized that gender was an important determinant in the current study, the analysis was stratified by gender. We adjusted for age; no adjustments were made for the other covariates because the purpose of this study was to detect target populations for future intervention programs. All *P*-values were two-sided, and statistical significance was set at *P* < .05. We used Stata/SE 16.1 (StataCorp, College Station, TX, USA) for all analyses.

## Results

Table 1 shows the characteristics of the study population. Data on a total of 27,036 participants (13,814 men and 13,222 women) were analyzed. We found that 43.0% of the participants reported that they had received the influenza vaccine during the current season. Regarding fear of COVID-19 transmission, 74.0% indicated that they felt fear. Overall, 37.5% of the participants expressed that they would like to get a COVID-19 vaccine if it becomes available.

**Table 1.** Characteristics of the study population.

|  | *n* (%) |
| --- | --- |
| Gender |  |
| Men | 13,814 (51.1) |
| Women | 13,222 (48.9) |
| Age (years) |  |
| 20–29 | 1,905 (7.0) |
| 30–39 | 4,858 (18.0) |
| 40–49 | 8,011 (29.6) |
| 50–59 | 9,012 (33.3) |
| 60–65 | 3,250 (12.0) |
| Education level |  |
| Junior high or high school | 7,321 (27.1) |
| Vocational school or college | 6,544 (24.2) |
| University or graduate school | 13,171 (48.7) |
| Marital status |  |
| Single | 9,164 (33.9) |
| Divorced or widowed | 2,843 (10.5) |
| Married | 15,029 (55.6) |
| Annual household income (JPY) |  |
| < 2,000,000 | 1,709 (6.3) |
| 2,000,000–3,999,999 | 5,548 (20.5) |
| 4,000,000–7,999,999 | 11,927 (44.1) |
| ≥ 8,000,000 | 7,852 (29.1) |
| Underlying disease |  |
| No | 17,526 (64.8) |
| Yes | 9,510 (35.2) |
| Smoking status |  |
| Never smoked | 14,587 (54.0) |
| Former smoker | 5,445 (20.1) |
| Current smoker | 7,004 (25.9) |
| Received the influenza vaccine during the current season |  |
| No | 15,411 (57.0) |
| Yes | 11,625 (43.0) |
| Fear of COVID-19 transmission |  |
| No | 7,019 (26.0) |
| Yes | 20,017 (74.0) |
| Willing to get the COVID-19 vaccine |  |
| No | 16,889 (62.5) |
| Yes | 10,147 (37.5) |

JPY: Japanese yen; COVID-19: coronavirus disease 2019

Table 2 displays the characteristics of the study population by willingness to get the COVID-19 vaccine and gender. A total of 5,780 men (41.8%) and 4,367 women (33.0%) were willing to get the COVID-19 vaccine. Relatively high percentages of participants were willing to get the COVID-19 vaccine among those aged 60–65 years (men: 48.0%; women: 41.1%), those who had an underlying disease (men: 47.8%; women: 37.0%), those who felt fear of COVID-19 transmission (men: 49.5%; women: 37.0%), and those who had received the influenza vaccine during the current season (men: 59.2%; women: 43.7%). These percentages were higher in men than in women.

**Table 2.**
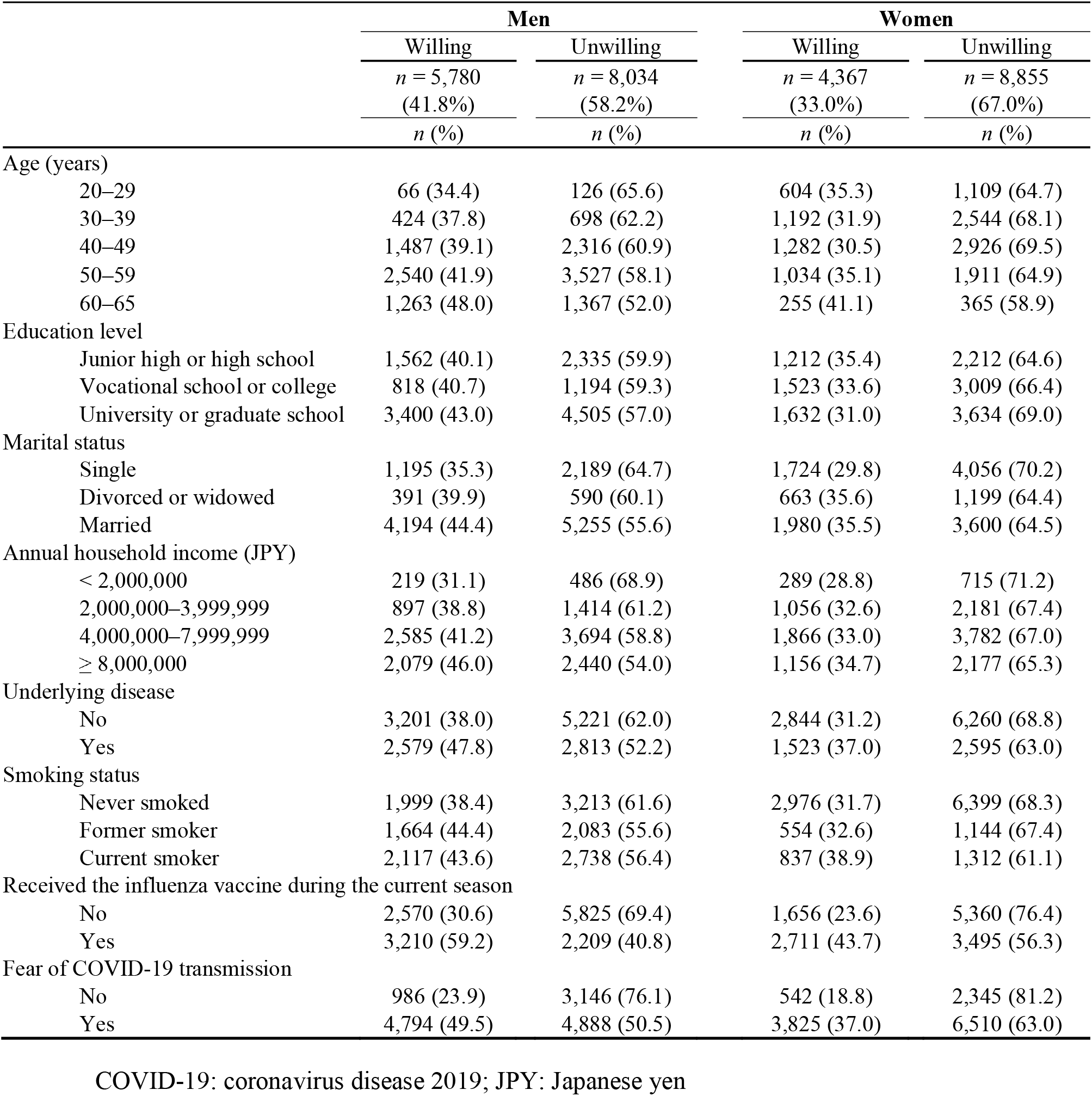
Characteristics of the study population by willingness to get the COVID-19 vaccine and gender.

|  | Men |  | Women |  |
| --- | --- | --- | --- | --- |
|  | Willing<br>*n* = 5,780<br>(41.8%)<br>*n* (%) | Unwilling<br>*n* = 8,034<br>(58.2%)<br>*n* (%) | Willing<br>*n* = 4,367<br>(33.0%)<br>*n* (%) | Unwilling<br>*n* = 8,855<br>(67.0%)<br>*n* (%) |
| Age (years) |  |  |  |  |
| 20–29 | 66 (34.4) | 126 (65.6) | 604 (35.3) | 1,109 (64.7) |
| 30–39 | 424 (37.8) | 698 (62.2) | 1,192 (31.9) | 2,544 (68.1) |
| 40–49 | 1,487 (39.1) | 2,316 (60.9) | 1,282 (30.5) | 2,926 (69.5) |
| 50–59 | 2,540 (41.9) | 3,527 (58.1) | 1,034 (35.1) | 1,911 (64.9) |
| 60–65 | 1,263 (48.0) | 1,367 (52.0) | 255 (41.1) | 365 (58.9) |
| Education level |  |  |  |  |
| Junior high or high school | 1,562 (40.1) | 2,335 (59.9) | 1,212 (35.4) | 2,212 (64.6) |
| Vocational school or college | 818 (40.7) | 1,194 (59.3) | 1,523 (33.6) | 3,009 (66.4) |
| University or graduate school | 3,400 (43.0) | 4,505 (57.0) | 1,632 (31.0) | 3,634 (69.0) |
| Marital status |  |  |  |  |
| Single | 1,195 (35.3) | 2,189 (64.7) | 1,724 (29.8) | 4,056 (70.2) |
| Divorced or widowed | 391 (39.9) | 590 (60.1) | 663 (35.6) | 1,199 (64.4) |
| Married | 4,194 (44.4) | 5,255 (55.6) | 1,980 (35.5) | 3,600 (64.5) |
| Annual household income (JPY) |  |  |  |  |
| < 2,000,000 | 219 (31.1) | 486 (68.9) | 289 (28.8) | 715 (71.2) |
| 2,000,000–3,999,999 | 897 (38.8) | 1,414 (61.2) | 1,056 (32.6) | 2,181 (67.4) |
| 4,000,000–7,999,999 | 2,585 (41.2) | 3,694 (58.8) | 1,866 (33.0) | 3,782 (67.0) |
| ≥ 8,000,000 | 2,079 (46.0) | 2,440 (54.0) | 1,156 (34.7) | 2,177 (65.3) |
| Underlying disease |  |  |  |  |
| No | 3,201 (38.0) | 5,221 (62.0) | 2,844 (31.2) | 6,260 (68.8) |
| Yes | 2,579 (47.8) | 2,813 (52.2) | 1,523 (37.0) | 2,595 (63.0) |
| Smoking status |  |  |  |  |
| Never smoked | 1,999 (38.4) | 3,213 (61.6) | 2,976 (31.7) | 6,399 (68.3) |
| Former smoker | 1,664 (44.4) | 2,083 (55.6) | 554 (32.6) | 1,144 (67.4) |
| Current smoker | 2,117 (43.6) | 2,738 (56.4) | 837 (38.9) | 1,312 (61.1) |
| Received the influenza vaccine during the current season |  |  |  |  |
| No | 2,570 (30.6) | 5,825 (69.4) | 1,656 (23.6) | 5,360 (76.4) |
| Yes | 3,210 (59.2) | 2,209 (40.8) | 2,711 (43.7) | 3,495 (56.3) |
| Fear of COVID-19 transmission |  |  |  |  |
| No | 986 (23.9) | 3,146 (76.1) | 542 (18.8) | 2,345 (81.2) |
| Yes | 4,794 (49.5) | 4,888 (50.5) | 3,825 (37.0) | 6,510 (63.0) |

COVID-19: coronavirus disease 2019; JPY: Japanese yen

Table 3 presents the factors associated with willingness to get the COVID-19 vaccine by gender. Men aged 50–59 years (aOR: 1.38; 95% CI: 1.02–1.86) and those aged 60–65 years (aOR: 1.76; 95% CI: 1.30–2.40) were more willing to get the COVID-19 vaccine than were men aged 20–29 years. Conversely, women aged 30–39 years (aOR: 0.86; 95% CI: 0.76–0.97) and those aged 40–49 years (aOR: 0.80; 95% CI: 0.71–0.91) were less willing to get the vaccine than were women aged 20–29 years. Regarding education level, compared with having a junior high or high school education, having a university or graduate school education was a facilitating factor for willingness to get the COVID-19 vaccine for men (aOR: 1.13; 95% CI: 1.04–1.22) but a barrier for women (aOR: 0.83; 95% CI: 0.75–0.91).

**Table 3.**
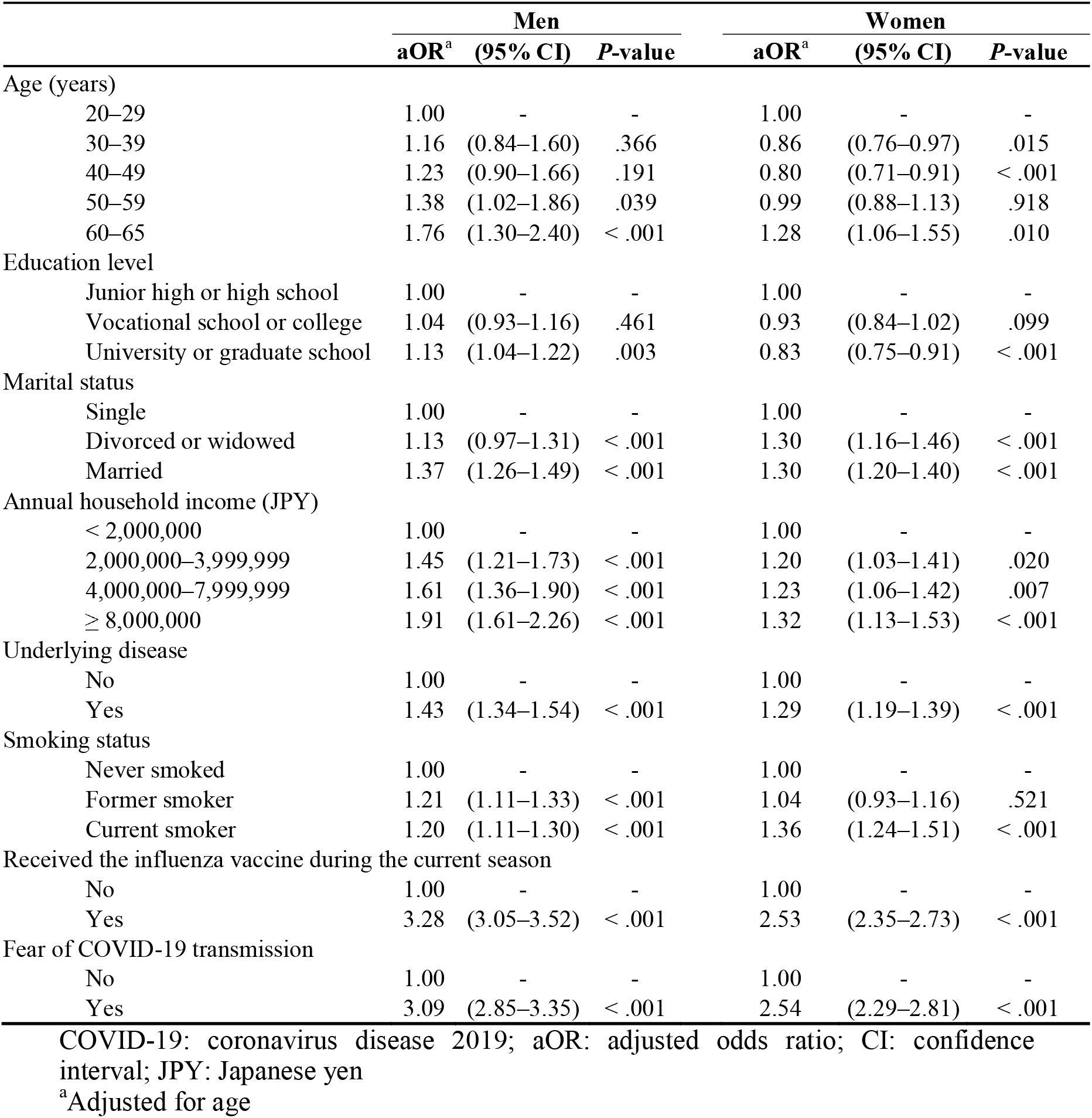
Factors associated with willingness to get the COVID-19 vaccine by gender.

For both men and women, a significantly higher likelihood of willingness to get the COVID-19 vaccine was observed among those who were married (men—aOR: 1.37; 95% CI: 1.26–1.49; women—aOR: 1.30; 95% CI: 1.20–1.40), those with a higher annual household income (men—aOR: 1.91; 95% CI: 1.61–2.26; women—aOR: 1.32; 95% CI: 1.13–1.53), those with an underlying disease (men—aOR: 1.43; 95% CI: 1.34– 1.54; women—aOR: 1.29; 95% CI: 1.19–1.39), those who currently smoked (men— aOR: 1.20; 95% CI: 1.11–1.30; women—aOR: 1.36; 95% CI: 1.24–1.51), those who had been vaccinated against influenza during the current season (men—aOR: 3.28; 95% CI: 3.05–3.52; women—aOR: 2.53; 95% CI: 2.35–2.73), and those with a fear of COVID-19 transmission (men—aOR: 3.09; 95% CI: 2.85–3.35; women—aOR: 2.54; 95% CI: 2.29–2.81).

## Discussion

This study provides evidence on gender differences in the determinants of willingness to get the COVID-19 vaccine. In the present study, the percentage of people who were willing to get the COVID-19 vaccine was lower among women than among men (33.0% vs. 41.8%). This trend was similar to the findings of previous studies in developed countries.^14, 19, 24, 25^ Notably, we found a gender gap in the associations of age and education level with willingness to get the COVID-19 vaccine: men who were older or had a higher level of education were more willing to get the vaccine, whereas women aged 30–49 years and those with a higher level of education were less willing to get the vaccine. Other factors showed similar trends for men and women: for both men and women, a higher likelihood of being willing to get the COVID-19 vaccine was observed in married participants, those with a higher annual household income, those who had an underlying disease, those who currently smoked, those who had been vaccinated against influenza during the current season, and those with a fear of COVID-19 transmission. These findings give important insight into identifying target groups for improving COVID-19 vaccination intentions, especially among women.

The most notable finding of the present study was a gender gap in the associations of age and education with willingness to get the COVID-19 vaccine. As mentioned above, men who were older or had a higher level of education were more willing to get the vaccine, whereas women aged 30–49 years and those with a higher level of education were less willing to get the vaccine. For men, this finding is consistent with previous studies; older age and higher level of education are known facilitators for health protection, which should correlate with COVID-19 vaccination intentions.^24, 26, 27^ Conversely, the current results for women vary from the findings presented in these studies. This difference may be explained by a higher level of concern about adverse effects of the vaccine among women aged 30–49 years and among those with a relatively high level of education in Japan during the current study period. We considered that recent reporting on adverse effects following receipt of the human papillomavirus vaccine in major Japanese newspapers may have had a negative impact on the vaccination intentions of these groups.^28^ These findings may imply the need for intervention in this population regarding COVID-19 vaccination.

The current study found that influenza vaccination status and fear of COVID-19 transmission were strongly associated with willingness to get the COVID-19 vaccine among both men and women. These results are in line with previous studies.^16, 17, 29, 30^ In the case of the 2009 influenza pandemic, perceived risk of infection translated into preventive behaviors, including vaccination.^31^ Similarly, the experience of receiving a vaccine have been shown to increase an individual’s confidence in the efficacy and safety of other vaccines.^29, 30^ Nevertheless, working-age people have fewer opportunities to receive the influenza vaccine compared with older adults because of the former group’s busy work schedules.^32^ These findings suggest that providing education about COVID-19 and influenza vaccination in the workplace may be an effective strategy to increase COVID-19 vaccine uptake.

In the current study, a higher likelihood of being willing to get the COVID-19 vaccine was observed among those who were married, those with a higher annual household income, those with an underlying disease, and those who currently smoked. Vulnerable populations, such as people with low socioeconomic status, are often found to have relatively poor health and to need support regarding their willingness to be vaccinated.^16^ In addition, a previous study on influenza vaccination suggests that single people are less willing to be vaccinated compared with their married counterparts because single people are not at risk of transmission from coresident family members.^11^ Our finding regarding the presence of underlying disease may be related to the fear of COVID-19 transmission because underlying disease a known risk factor for clinical severity of COVID-19 infection.^24^ Only our finding on tobacco use was inconsistent with previous studies, which have found that smokers often tend to refuse vaccines regardless of the risk of clinical worsening associated with smoking.^10, 11, 33^ Further studies should focus on confirming the relationship between smoking and COVID-19 vaccination intention.

Overall, we found that 37.5% of the working-age population in Japan was willing to get the COVID-19 vaccine. This percentage is not high enough to achieve herd immunity through the vaccination program.^5^ This acceptance rate is lower than that in other countries, such as China (91.3%), France (58.9%), the US (56.9%), and Russia (54.9%).^6^ Furthermore, the acceptance rate is lower than the rate reported in a previous study in Japan (65.7%) that was conducted in September 2020.^19^ One possible reason for the low vaccination intention in the current study may be tied to the research period.^15^ Our study was conducted in December 2020, right after the start of the COVID-19 vaccination program in the UK and the US, and the media coverage of adverse effects was exaggerated.^34, 35^ Another reason is that people of working age have been found to be less willing to get the COVID-19 vaccine compared with older adults,^19^ which would affect the present results. These findings have important implications for vaccination intentions, although vaccination acceptance may change after the vaccine is available for administration among the working-age population.

The main strengths of this study are the large sample size and the use of a sample from throughout Japan. A limitation of this study is that we recruited study participants from an Internet research company’s list of panelists. A previous study on the anti-vaccine movement reported that anti-vaccine messages were more prevalent on the Internet than in other sources.^36^ Therefore, the participants in the current study may have been particularly likely to access anti-vaccine websites, and this should be taken into account when interpreting the results of our study. Additionally, we conducted the current study before the administration of the COVID-19 vaccination program in Japan; therefore, we could not provide participants with detailed information, such as the vaccination schedule, which may have affected their willingness to get the vaccine.

In conclusion, the current study revealed a gender gap in the associations of age and education level with willingness to get the COVID-19 vaccine. In particular, women aged 30–49 years and those with a higher level of education were less willing to get the vaccine. These findings may imply the need for intervention for this population regarding COVID-19 vaccination. Providing education about COVID-19 and influenza vaccination in the workplace may be an effective strategy to increase COVID-19 vaccine uptake.

## Data Availability

No additional data are available.

## Acknowledgments

We thank the current members of the CORoNaWork Project (listed in alphabetical order): Professor Yoshihisa Fujino (present chairperson of the study group), Dr. Hajime Ando, Professor Hisashi Eguchi, Dr. Kazunori Ikegami, Dr. Tomohiro Ishimaru, Dr. Arisa Harada, Dr. Ayako Hino, Dr. Kyoko Kitagawa, Dr. Kosuke Mafune, Professor Shinya Matsuda, Dr. Ryutaro Matsugaki, Professor Koji Mori, Dr. Keiji Muramatsu, Dr. Masako Nagata, Dr. Tomohisa Nagata, Ms. Ning Liu, Professor Akira Ogami, Dr. Rie Tanaka, Dr. Seiishiro Tateishi, Dr. Kei Tokutsu and Professor Mayumi Tsuji. All members are affiliated with the University of Occupational and Environmental Health, Japan.

